# A SARS-CoV-2 serological assay to determine the presence of blocking antibodies that compete for human ACE2 binding

**DOI:** 10.1101/2020.05.27.20114652

**Authors:** James R. Byrnes, Xin X. Zhou, Irene Lui, Susanna K. Elledge, Jeff E. Glasgow, Shion A. Lim, Rita Loudermilk, Charles Y. Chiu, Michael R. Wilson, Kevin K. Leung, James A. Wells

## Abstract

As SARS-CoV-2 continues to spread around the world, there is an urgent need for new assay formats to characterize the humoral response to infection. Convalescent serum is being used for treatment and for isolation of patient-derived antibodies. However, currently there is not a simple means to estimate serum bulk neutralizing capability. Here we present an efficient competitive serological assay that can simultaneously determine an individual’s seropositivity against the SARS-CoV-2 Spike protein and estimate the neutralizing capacity of anti-Spike antibodies to block interaction with the human angiotensin converting enzyme 2 (ACE2) required for viral entry. In this ELISA-based assay, we present natively-folded viral Spike protein receptor binding domain (RBD)-containing antigens via avidin-biotin interactions. Sera are then supplemented with soluble ACE2-Fc to compete for RBD-binding serum antibodies, and antibody binding quantified. Comparison of signal from untreated serum and ACE2-Fc-treated serum reveals the presence of antibodies that compete with ACE2 for RBD binding, as evidenced by loss of signal with ACE2-Fc treatment. In our test cohort of nine convalescent SARS-CoV-2 patients, we found all patients had developed anti-RBD antibodies targeting the epitope responsible for ACE2 engagement. This assay provides a simple and high-throughput method to screen patient sera for potentially neutralizing anti-Spike antibodies to enable identification of candidate sera for therapeutic use.

## INTRODUCTION

The emergence of the pathogenic SARS-CoV-2 coronavirus in humans, and the respiratory disease associated with infection, coronavirus disease 2019 (COVID-19), has placed a significant public health burden on countries around the world. SARS-CoV-2 is closely related to the pathogenic SARS-CoV-1 that emerged in the early 2000s.^1^ Both viruses infect bronchial epithelial cells and type II pneumocytes, and enter human cells via the host receptor angiotensin-converting enzyme 2 (ACE2).^2,3^ Viral entry is dependent on a binding interaction between the receptor binding domain (RBD) of the viral Spike protein and ACE2 on the cell surface.^4,5^ Given the crucial role of RBD binding to ACE2 in viral entry, disrupting this interaction has emerged as a promising target for first-generation biologics to provide passive immunity, either with anti-Spike antibodies^6^ or ACE2 constructs^7-9^. As more patients recover from SARS-CoV-2 infection, there is a great need for serology assays to examine the humoral response to infection and vaccination, as well as to determine how patient-derived antibodies interact with the Spike protein.

Although direct detection of viral proteins or polymerase chain reaction testing is key to diagnosing the early stages of SARS-CoV-2 infection, serological assays detecting anti-SARS-CoV-2 antibodies are vital tools for monitoring how a patient’s anti-viral response evolves after the period of acute infection ends.^10^ Serological assays can take many forms, including enzyme-linked immunosorbent assays (ELISA)^11^, viral neutralization assays, and rapid lateral flow assays.^10^ Neutralization assays with serum necessitate culture of either live or pseudovirus, and the market has been flooded with rapid lateral flow diagnostic tests that provide heterogeneous results^12^ and are difficult to quantify.

ELISA-based tests are not as rapid, but they provide quantitative results and are easily adapted to test a variety of conditions and experimental designs. Such variations in design can provide greater detail regarding the profile of a patient’s antibody response to SARS-CoV-2 and how these antibodies interact with viral antigens. One clear application of a modified ELISA-type serology assay is to rapidly screen for the presence of patient antibodies that compete with ACE2 for RBD binding, and therefore may disrupt RBD binding to ACE2. Improved understanding of the prevalence of these antibodies in patient sera will inform both therapeutic and vaccine design efforts and allow for correlations between clinical outcome and the presence of potentially neutralizing antibodies. Furthermore, recent therapeutic use of convalescent sera shows that while safe, the efficacy of this treatment still remains unclear.^13,14^ A serological assay capable of simultaneously screening convalescent sera for potentially neutralizing antibodies would offer clinicians a facile way to screen for high-titer neutralizing convalescent sera that may be effective in treating acutely ill individuals and for speeding population-based studies to understand what proportion of the exposed population might have a degree of protective immunity.

Herein, we report the development of a simple competitive serological assay using biotinylated Spike protein antigens and a dimeric ACE2-Fc fusion construct. Use of the avidin-biotin interaction to coat plates with biotinylated antigen versus simple adsorption permits the presentation of natively-folded protein for serum antibody capture. Our assay is analogous to the widely-used RBD ELISA assay first reported by Amanat et al.^15^ and later expanded by Stadlbauer et al.^11^ However, we show here that the addition of ACE2-Fc competitor to the sera allows us to also examine potentially neutralizing antibodies that block ACE2-RBD interactions. The competition reactions are performed on the same plate, using the same detection protocol to enable rapid, reproducible characterization of a patient’s anti-Spike antibody profile and whether the patient has generated antibodies that compete with ACE2 for RBD binding. Importantly, when used in conjunction with anti-Spike antibodies with known binding epitopes, this assay format can be further adapted to reveal Spike epitopes targeted by patient antibodies at higher resolution, providing important insights for the development of biologic therapies and vaccines.

## RESULTS

### Biotinylated SARS-CoV-2 spike protein antigen constructs offer multiple formats to detect anti-Spike antibodies in convalescent patient sera

Recently, we reported the high-yield expression of a number of biotinylated SARS-CoV-2 Spike protein antigen and ACE2 formats that have broad utility for SARS-CoV-2 research.^16^ Given the importance of serological testing to identify not only patients that have recovered from SARS-CoV-2 infection, but also the need to characterize the properties of anti-Spike antibodies generated by these patients, we developed a serological assay using these biotinylated constructs (Table 1, Figure 1A, Supplemental Figure 1). Briefly, plates are first coated with NeutrAvidin followed by incubation with biotinylated antigen similar to our previously reported assays to characterize Fab-phage binding as part of our phage display recombinant antibody generation pipeline^17^. Plates are then blocked using 3% nonfat milk and incubated with serum diluted in 1% nonfat milk (Figure 1A). The use of a standard anti-IgG-horseradish peroxidase (HRP) as a detection reagent is precluded by our incorporation of a human Fc region into some of our Spike antigen constructs for dimeric presentation of the RBD. Thus, our pilot studies first focused on testing multiple alternative detection reagents. In all studies presented here, convalescent patients had prior RT-PCR-confirmed SARS-CoV-2 infection, and sera were collected 14 or more days following resolution of COVID-19 symptoms. Healthy control serum was collected before the emergence of SARS-CoV-2. We found that anti-Fab-HRP, anti-IgM-HRP, and Protein L-HRP all offered robust detection of anti-Spike antibodies in patient sera (Supplemental Figure 2). Protein L-HRP was predominantly used in this study as it will detect any antibody subtype containing a kappa-light chain (e.g. IgG, IgM, IgA). We observed slight reactivity of Protein L-HRP to Fc-containing antigens in buffer-treated wells (Supplemental Figure 2C), so in subsequent assays the background signal in buffer-treated wells was subtracted from the sample signal. Further pilot studies to optimize our workflow revealed that heat inactivation of patient serum (56°C, 60 minutes) did not significantly reduce signal (P=0.4877, Supplemental Figure 3), consistent with previous reports^15^. Additionally, these early experiments showed that coating with as low as 20 nM RBD-biotin still provided robust detection of anti-RBD patient antibodies (Supplemental Figure 4). In summary, we converged on an optimal assay condition utilizing a 20 nM antigen coating concentration, 50-fold diluted, heat-inactivated sera, and Protein-L-HRP as a detection reagent.

**Table 1.** Protein constructs used in competitive SARS-CoV-2 serology assay.

| **Construct** | **Category** | **Description** |
| --- | --- | --- |
| RBD-biotin | Antigen | Monomeric SARS-CoV-2 RBD (Spike residues 328-533) |
| FL Spike-biotin* | Antigen | Full-length spike ectodomain containing S1, S2, and RBD domains, trimer |
| RBD-hFc | Antigen | Fusion of SARS-CoV-2 RBD to a human IgG1 Fc, dimer |
| RBD-mFc | Antigen | Fusion of SARS-CoV-2 RBD to a mouse IgG2a Fc, dimer |
| ACE2-Fc** | Competitor | Fusion of human ACE2 extracellular domain to a human IgG1 Fc via a tobacco etch virus (TEV) protease-cleavable linker, dimer |
| ACE2 monomer | Competitor | Extracellular domain of ACE2 (residues 1-614), monomer generated by TEV protease digestion of ACE2-Fc construct<sup>16</sup> |

\*Biotinylated version of FL Spike construct designed by Amanat et al.<sup>15</sup>
\*\*all proteins are biotinylated via an Avi-tag aside from ACE2-Fc

**Figure 1.**
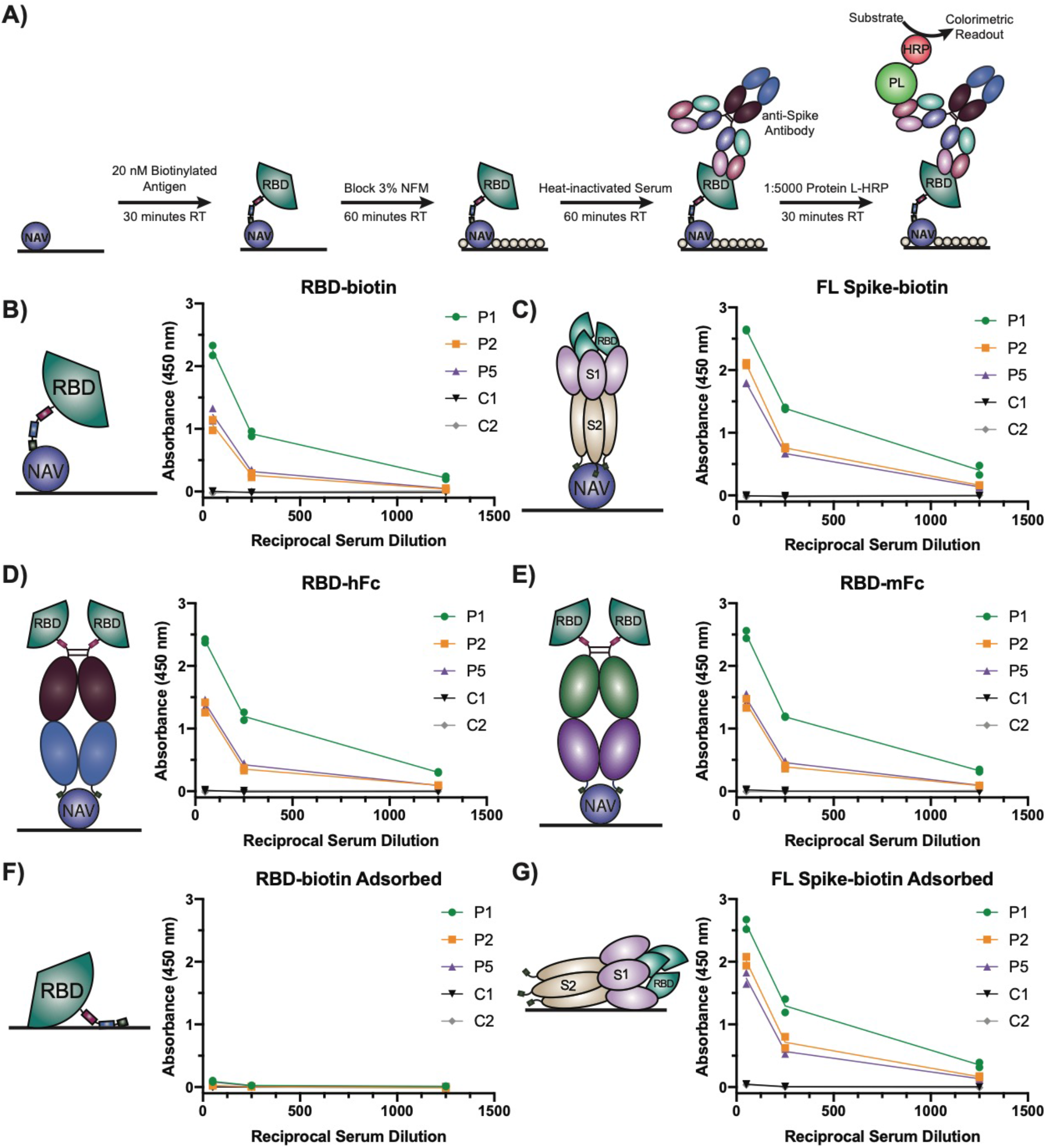
Native presentation of SARS-CoV-2 Spike protein antigens effectively detect anti-Spike antibodies in convalescent patient sera. (A) Schematic of NeutrAvidin:biotinylated antigen serology ELISA setup and detection strategy using Protein L-HRP (PL-HRP). ELISA results for the indicated antigens presented via NeutrAvidin (NAV, B-E) or passively adsorbed to the plate (F-G). Sera from three patients (P1, P2, P5) and two healthy controls (C1, C2) were serially diluted five-fold from an initial 50-fold dilution. Antigen coating solutions were 20 nM. Each sample was run with two technical replicates on the plate. Dots indicate the mean signal of technical replicates from each of two (N=2) independent experiments. NFM=nonfat milk.

To profile the efficacy of our various antigens to directly detect anti-Spike antibodies in patient sera, we performed a head-to-head comparison of all antigen constructs listed in Table 1. All biotinylated constructs were immobilized on NeutrAvidin-coated plates and effectively captured anti-Spike antibodies from three patient sera, whereas sera from two healthy controls were not reactive (Figure 1B-E). We observed a dose-dependent decrease in signal with increasing serum dilution for all antigens. All three patients tested exhibited strong reactivity to both RBD-biotin and biotinylated full-length (FL) Spike protein. Of note, the use of a human (RBD-hFc) or mouse Fc-fusion (RBD-mFc) did not affect signal strength (Figure 1D-E). There also did not seem to be a clear benefit of monomeric (RBD-biotin, Figure 1B) versus dimeric (RBD-hFc, RBD-mFc, Figure 1D-E) presentation of the RBD, aside from slightly higher signal at a 1:250 serum dilution.

Interestingly, passive adsorption of RBD-biotin to the plate instead of utilizing the interaction with NeutrAvidin resulted in loss of signal (Figure 1F). Conversely, adsorption of FL Spike-biotin did not affect signal (Figure 1G). To further probe this observation, we tested adsorption of both 20 and 100 nM RBD-biotin and found that adsorption with a low concentration of RBD-biotin (20 nM) and detection with Protein L-HRP were not compatible, underscoring the importance of presenting antigens via NeutrAvidin in our experimental design (Supplemental Figure 5). However, adsorption at a higher RBD-biotin concentration (100 nM) yielded signal with Protein L-HRP. Furthermore, detection with anti-human IgG in a format analogous to previously reported assays^11,15^ yielded robust signal when 100 nM RBD-biotin was adsorbed (Supplemental Figure 5D), indicating RBD-biotin can also be used in an adsorption format.

Not surprisingly, we observed higher signal at all serum dilutions with FL Spike-biotin (413 kDa) than RBD-biotin (28.5 kDa). However, the signal increase was less than two-fold, while the size difference between these proteins by molecular weight is >14-fold. This observation suggests that a large proportion of anti-Spike antibodies that patients develop specifically target the RBD domain.

### ACE2-Fc competes with antibodies in a majority of patient sera tested

We next adapted our assay to incorporate an ACE2 competition condition where patient antibodies compete in the presence of ACE2 for binding to Spike antigen on the ELISA plate (Figure 2A). We first tested the monomeric ACE2 and observed a modest, but consistent reduction in bound antibody signal across the four patient samples tested (Supplemental Figure 6). We have previously shown dimeric ACE2-Fc binds ~4-fold tighter to the monomeric Spike antigen.^16^ Therefore, we postulated the improved affinity and potential avidity afforded with this dimeric construct may allow greater competition with patient antibodies. Indeed, we observed a much greater decrease in RBD-binding of patient antibodies when serum was supplemented with 100 nM ACE2-Fc (Figure 2B), a concentration of ACE2-Fc we found to saturate RBD on the plate (Supplemental Figure 7). Pre-treating the antigen-coated plate with ACE2-Fc prior to adding serum produced slightly higher signal compared to when ACE2-Fc was added to serum. This suggests that pre-treatment with ACE2-Fc allows for some dissociation of ACE2-Fc during serum incubation, and consequently increased patient antibody binding (Supplemental Figure 8). Therefore, we chose to supplement sera with ACE2-Fc to allow simultaneous competition with the patient antibodies to bind immobilized Spike antigen.

**Figure 2.**
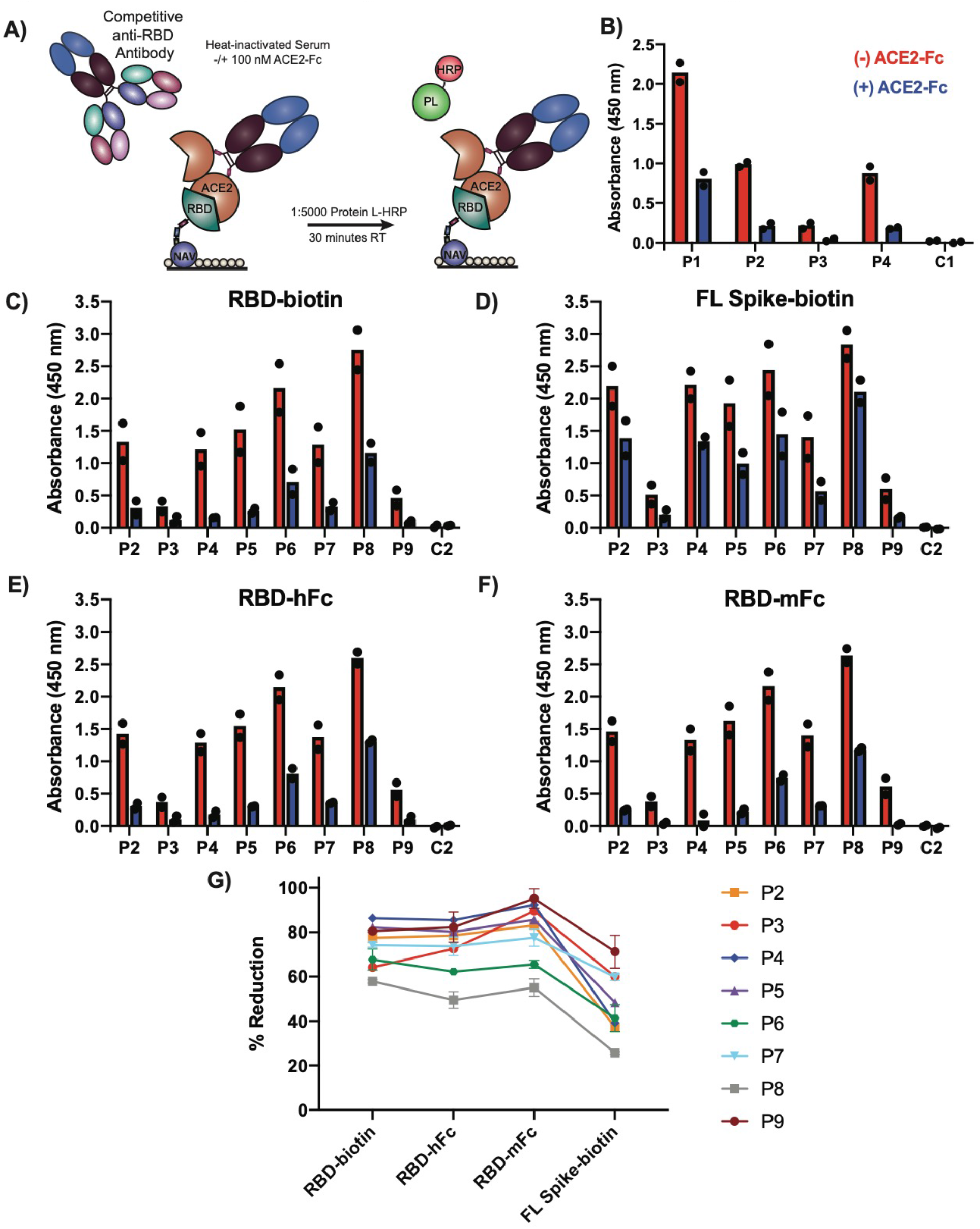
ACE2-Fc competes with patient antibodies for RBD binding. (A) Schematic of ACE2-Fc competitive serology ELISA setup. (B) Competition ELISA (100 nM ACE2-Fc) results from four patients (P1-P4) and one healthy control (C1) using RBD-biotin as the capture antigen. (C-F) Competition ELISA (100 nM ACE2-Fc) results using the indicated antigens for eight patients (P2-P9) and one healthy control (C2). All sera were diluted 1:50 for analysis. Each sample was run with two technical replicates on the plate. Dots indicate mean signal of technical replicates from two (N=2) independent experiments. Bar shows the mean of these two experiments. (G) Percent reduction in signal seen on inclusion of 100 nM ACE2-Fc in serum dilution. Dots represent mean −/+ standard deviation of N=2 experiments.

To determine the patient-to-patient variability in our ACE2-Fc competition serology assay, and to test our various antigen formats in this competition mode, we expanded our efforts to test a cohort of eight convalescent patient sera and a healthy control using the antigens presented in Table 1. We observed a ten-fold variation in the overall anti-Spike signal between patients, and trends were consistent between antigens (Figure 2C-F). When specifically examining the RBD-alone containing antigens (RBD-biotin, RBD-hFc, RBD-mFc, Figure 2C, E-F), all patients exhibited a variable, but substantial degree of signal decrease when ACE2-Fc was added to the serum (50-95%, Figure 2G). This finding suggests that the patients in this small cohort have all generated anti-RBD antibodies that bind at the RBD epitope responsible for binding ACE2, and are therefore potentially neutralizing.

Interestingly, when FL Spike-biotin was used as the antigen (Figure 2D), the signal for both direct detection and ACE2-Fc competition were largely elevated relative to the RBD-alone containing antigens. The average percent decrease in signal with ACE2-Fc competition was also lower than with the RBD-alone containing antigens (Figure 2G). These observations likely represent anti-Spike antibodies that bind outside of the RBD that are unaffected by ACE2-Fc competition. Taken together, these results demonstrate the utility of our ACE2-Fc competition assay for the simultaneous determination of both baseline serum reactivity to Spike antigens as well as whether a serum sample contains antibodies that compete for the ACE2-binding epitope on the RBD.

## DISCUSSION

As the SARS-CoV-2 pandemic escalates, there is a continued need for assays to profile patient responses to infection, especially with respect to the anti-viral antibodies generated, and whether or not a patient has acquired immunity against SARS-CoV-2. Here, we designed and employed an assay to identify potentially neutralizing antibodies in convalescent patient sera that bind at the ACE2/Spike RBD interface. Using a variety of biotinylated Spike antigens and presentation of natively-folded protein via avidin-biotin interactions, we developed another ELISA format for directly measuring patient seropositivity to the SARS-CoV-2 Spike protein. Competition with ACE2-Fc clearly revealed the presence of potential neutralizing antibodies that bind the RBD in all patients tested. This new assay represents a high-throughput and simple means of providing additional resolution of the patient antibody response to SARS-CoV-2 infection and further justifies efforts to design therapies and vaccines that block binding of the Spike RBD to ACE2.

Our cohort of convalescent patients was small, but several interesting trends can be observed in our results. There was remarkable consistency in detection using our various antigens. We were surprised to find that presentation of monomeric RBD-biotin versus dimeric RBD found in RBD-hFc and RBD-mFc showed little difference in signal strength. This may be a result of using tetrameric NeutrAvidin to present RBD-biotin, which would mimic an avidity effect. Another interesting trend is the varying degree of anti-Spike antibody seropositivity between patients. This variation is consistent with previous observations of heterogeneous responses of patient sera in anti-Spike serology assays.^15,18-21^ We also saw anti-Spike seroconversion in all of our patient samples, and reactivity to FL Spike-biotin was higher than that to antigens only containing the RBD, consistent with earlier observations.^15^ Taken together, these data demonstrate that our biotinylated antigen constructs can be effectively presented using immobilized avidin, and offer another option for serologic screening of individuals for anti-SARS-CoV-2 immunity.

An important advance presented here is the implementation of a straightforward means to assess the global capacity of a patient’s serum antibodies to compete with ACE2 for RBD binding. By simply adding ACE2-Fc to the serum dilution buffer, we can modify our direct-detection ELISA to reveal the presence of antibodies that bind at a potentially neutralizing RBD epitope in the ACE2/RBD interface. We found that all of the patients in our cohort had antibodies that bound RBD at or near this interface, as indicated by reductions in signal strength in the competition mode of our ELISA. These findings indicate that not only is this ACE2-binding surface of the RBD highly immunogenic, but also suggests that recovered COVID-19 patients develop antibodies against this potentially neutralizing epitope. In the context of previous findings that SARS-CoV-1 neutralizing antibodies bind the Spike RBD and block ACE2 binding^22,23^, our data suggest that this premise is also true for SARS-CoV-2. Furthermore, our results indicate that in most of the patients tested here, a majority of the anti-RBD antibodies bind at the ACE2 binding site on the RBD.

To our knowledge, only one other SARS-CoV-2 study has examined the ability of serum antibodies to compete with ACE2 for RBD binding.^18^ Interestingly, in contrast to our findings, only 3 out of 26 patient tested positive for ACE2-competitive antibodies. These divergent results are possibly a consequence of differing assay designs and competitor concentrations, or differing selection criteria of patient cohorts. We found in our experiments that ACE2 monomer could not efficiently compete with patient antibodies for binding, which underscores the importance of using a strong, bivalent binder to block the Spike-patient antibody interaction in such a competitive serology assay. We next plan to apply our competition ELISA in an expanded patient cohort with concurrent viral neutralization assays to further understand the prevalence of ACE2-competitive antibodies in SARS-CoV-2 convalescent patient sera and correlate these findings with clinical outcomes. These studies will build on our intriguing early results and help improve our understanding of the role binding epitope plays in SARS-CoV-2 neutralization.

Although our use of Protein L to detect patient antibodies allows us to simultaneously detect IgG, IgM, and IgA, it does not detect lambda light chain-containing antibodies. Testing of additional detection reagents and designs of avid ACE2 constructs that lack an Fc may identify alternatives that will allow simultaneous capture of antibody subtypes with either light chain. Another consideration is that the strength of ACE2 competition may not correlate with neutralization efficiency, and that our assay does not reveal if patients have sufficient titers of these competitive antibodies to provide effective neutralization. This underscores the need for additional experiments to perform the competition assay and serum neutralization experiments in parallel. Lastly, it is possible that an antibody could bind outside of the ACE2 binding epitope on the RBD but still prevent ACE2 binding via allosteric effects. Given the relatively small size of the RBD, this may be more relevant for the FL Spike protein, but if such an antibody is identified, testing it under our competition conditions would reveal if this binding would be detected in our assay design.

In conclusion, we developed a simple ELISA-based ACE2 competition serology assay to identify individuals with anti-SARS-CoV-2 antibodies against the ACE2-binding epitope on the Spike RBD. Addition of ACE2-Fc to serum is straightforward, does not significantly lengthen the ELISA protocol, and could potentially be an added step into other assay formats, such as lateral flow. This high-throughput assay could be used to pre-screen convalescent serum for bulk quality of potentially neutralizing antibodies and also to better inform recovered COVID-19 patients and their physicians whether their SARS-CoV-2 exposure has resulted in protective immunity or not. This design also has the potential to screen engineered Spike-binding proteins with potential therapeutic utility to determine if these constructs bind similar epitopes to antibodies generated in convalescent patients. Future studies will focus on increasing the number of patients screened as well as correlating results of the competition assay with clinical outcomes and SARS-CoV-2 neutralization experiments.

## MATERIALS AND METHODS

### Antigen Generation

All antigens and ACE2 constructs were produced as previously described.^16^ RBD-mFc was generated by subcloning the RBD DNA sequence into a vector containing a C-terminal mIgG2a-Fc with an Avi tag. The sequence maps for all plasmids are available upon request. Briefly, proteins were expressed in Expi293 cells co-expressing BirA using the Expifectamine Expression System Kit in accordance with the manufacturer’s recommended protocol (Thermo Fisher Scientific). Biotinylated proteins were then purified using either Ni-NTA (RBD-biotin, FL Spike) or Protein A chromatography (RBD-hFc, RBD-mFc) and buffer exchanged into phosphate-buffered saline (PBS) for storage at −80°C. Protein purity was assessed using SDS-PAGE and size exclusion chromatography. Biotinylation was confirmed by NeutrAvidin (Thermo Fisher Scientific) shift assay.

### Patient Samples

All serum samples were obtained via antecubital venipuncture and collected into BD Vacutainer serum collection tubes in using protocols approved by the UCSF Institutional Review Board and in accordance with the Declaration of Helsinki. All patients had a positive clinical nasopharyngeal RT-PCR test to document SARS-CoV-2 infection. At the time of their blood draw, more than 14 days had elapsed since their COVID-19 respiratory and constitutional symptoms had resolved. De-identified serum was aliquoted, flash frozen in liquid nitrogen, and stored at −80°C in single-use aliquots. Control samples were from healthy individuals obtained before the emergence of SARS-CoV-2. Heat-inactivation of sera was performed by incubating the samples at 56°C for 60 minutes.

### Competition ELISA Protocol

All assays were performed in 384-well Nunc Maxisorp flat-bottom plates (Thermo Fisher Scientific), and each sample was run in duplicate. First, plates were coated with 50 μL of 0.5 μg/mL NeutrAvidin or 20 μL of 20 nM antigen (for passively-adsorbed antigens) in PBS for 60 minutes at room temperature. For assays using 100 nM biotinylated antigen, 10 μg/mL NeutrAvidin was used. Plates were then washed 3X with PBS containing 0.05% Tween 20 (PBST), and were washed similarly between each of the following steps. Next, 20 μL of biotinylated antigens were added to NeutrAvidin-coated wells and allowed to bind for 30 minutes at room temperature. After washing, plates were then blocked for 60 minutes with 100 μl 3% nonfat milk (Lab Scientific) in PBST. Sera were diluted as indicated in 1% nonfat milk in PBST in the absence (direct detection) or presence (competition) of 100 nM ACE2-Fc and 20 μL of these dilutions were incubated in the plates for 60 minutes at room temperature. Plates were again washed, and antibodies bound to the coated antigens detected using 20 μL of anti-human Fab (Jackson ImmunoResearch Laboratories 109-036-097, [1:5000]), anti-human IgM (Sigma-Aldrich A6907, [1:3000]), anti-human IgG (Sigma-Aldrich A0170, [1:3000]), or Protein L (Thermo Fisher Scientific 32420, [1:5000]) as indicated for 30 minutes at room temperature. All detection reagents were conjugated to HRP. Following a final wash, plates were developed for 10 minutes at room temperature using 20 of 50/50 TMB/solution B (VWR International). Reactions were quenched with 20 μL 1 M phosphoric acid and absorbance measured at 450 nm using a Tecan Infinite M200 Pro spectrophotometer.

### Data Analysis and Statistics

Background from the raw ELISA signal was removed by first subtracting the signal measured in NeutrAvidin-alone coated wells or empty wells (passively adsorbed antigens). Next, the signal measured in antigen-coated wells incubated with 1% nonfat milk (direct detection) or 1% nonfat milk + 100 nM ACE2-Fc (competition) was subtracted from the signal in serum-treated wells. As there is some detectable reactivity of Protein L-HRP to Fc-containing antigens (RBD-hFc, RBD-mFc) and RBD-bound ACE2-Fc (competition mode), this buffer subtraction step is necessary. All graphing and statistical analysis was performed in GraphPad Prism (Version 8.4.2).

## Data Availability

All data presented are available upon request to the corresponding author.

## ACKNOWLEDGEMENTS

We acknowledge the members of the Wells Lab, especially those involved in our COVID-19 research program. We thank Dr. Peter Kim and Dr. Abigail Powell for helpful discussions. We also thank Dr. John Pak (Chan Zuckerberg Initiative Biohub) and Dr. Florian Krammer (Mt. Sinai Icahn School of Medicine) for providing the FL Spike plasmid for cloning a version containing an Avi tag for biotinylation. We thank Kanishka Koshal and Kelsey Zorn for their assistance with patient consenting and enrollment. We thank the patients for their participation in this study.

J.A.W. is grateful for funding from the Harry and Dianna Hind Endowed Professorship in Pharmaceutical Sciences and the Chan Zuckerberg Biohub that helped support this work. Postdoctoral Fellowship support included a National Institutes of Health National Cancer Institute F32 (5F32CA239417 to J.R.B.), Damon Runyon Postdoctoral Fellowship (to X.X.Z.), and a Merck Postdoctoral Research Fellowship from the Helen Hay Whitney Foundation (S.A.L.). The National Science Foundation Graduate Research Fellowship Program supported I.L. and S.K.E. Study enrollment and collection of patient and control sera were supported in part by National Institutes of Health grants R01-HL105704 (to C.Y.C.) from the National Heart, Lung, and Blood Institute, R33-129077 (to C.Y.C.) from the National Institute of Allergy and Infectious Diseases, and the Charles and Helen Schwab Foundation (to C.Y.C.). M.R.W. also thanks an endowment from the Rachleff Family for support.

## AUTHOR CONTRIBUTIONS

J.R.B. designed the research, performed experiments, and analyzed data. X.X.Z., I.L., J.E.G., S.K.E., S.A.L. and K.K.L. designed research and helped with protein design, expression, and purification. R.L., C.Y.C., and M.R.W. provided patient and control sera. J.A.W. supervised the research. J.R.B., X.X.Z, K.K.L., and J.A.W. co-wrote the manuscript, and all authors provided edits and approval of the final version.

---

**Supplemental Figure 1.**
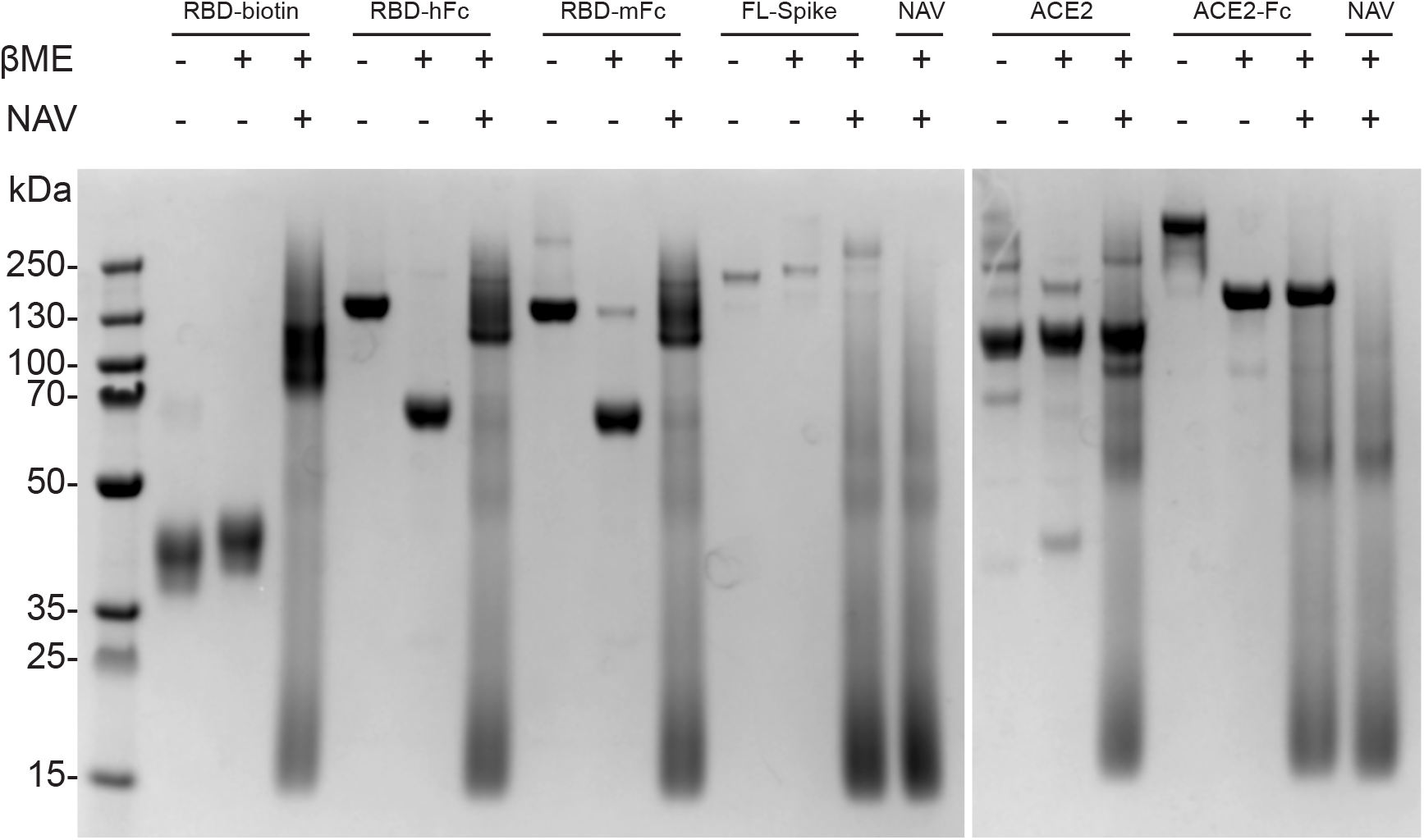
SDS-PAGE of proteins used in serology assays. All proteins were prepared as previously described^16^ and visualized with SDS-PAGE either in the presence or absence of β-mercaptoethanol and NeutrAvidin (NAV) as indicated.

**Supplemental Figure 2.**
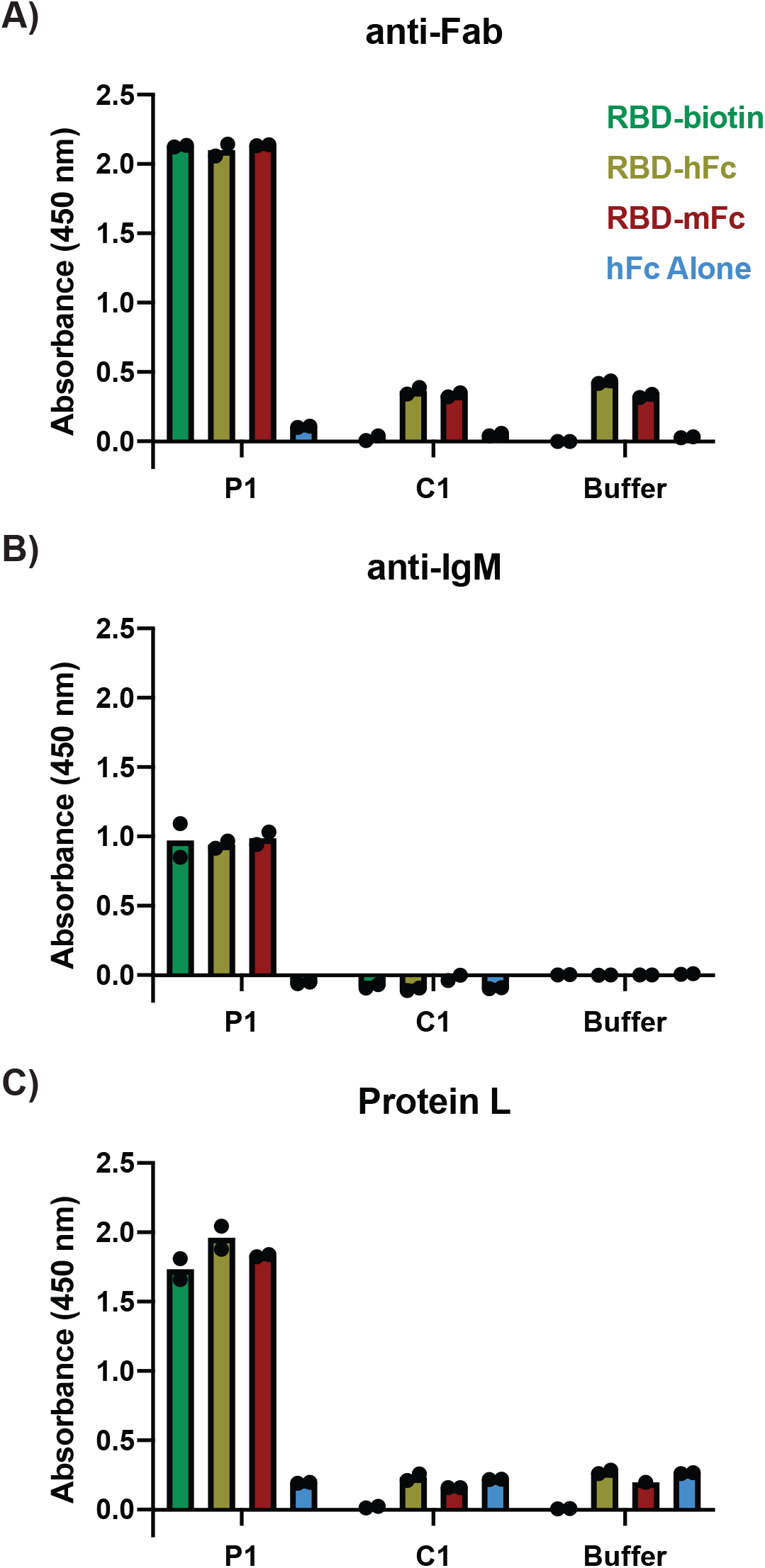
HRP-conjugated anti-Fab, anti-IgM, and Protein L are effective secondaries. ELISA results using the indicated antigens and detection with either (A) anti-Fab-HRP, (B) anti-IgM-HRP, or (C) Protein L-HRP secondaries for one patient and one healthy control. The buffer condition represents wells that received 1% nonfat milk in PBST instead of serum. Sera were diluted 1:50. Dots represent values of technical replicates, bars represent the mean of these replicates.

**Supplemental Figure 3.**
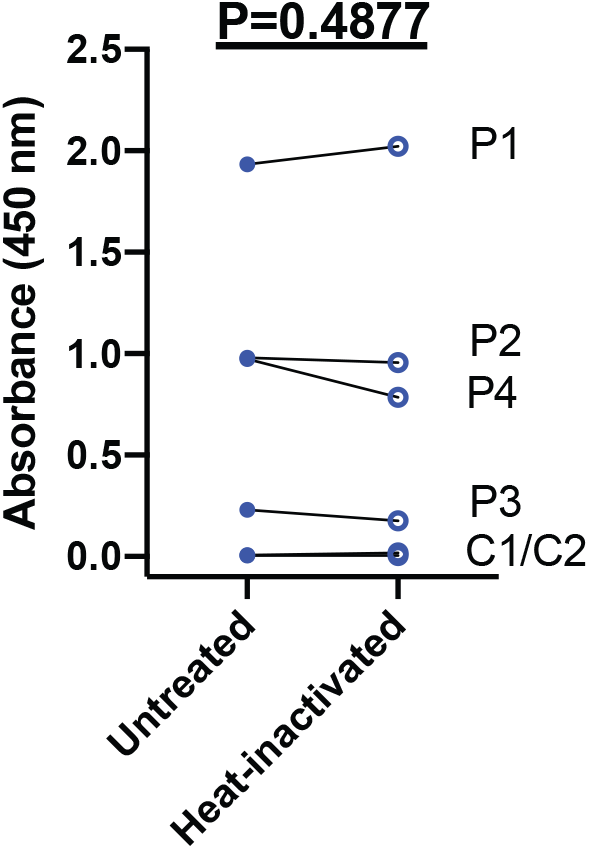
Heat-inactivation of sera at 56°C for 60 minutes does not affect detection with Protein L-HRP. ELISA results from wells coated with 20 nM RBD-biotin and incubated with either untreated or heat-inactivated sera from four patients and two healthy controls. Sera were diluted 1:50. Dots represent mean value of two technical replicates. A paired t-test was performed to determine statistical significance.

**Supplemental Figure 4.**
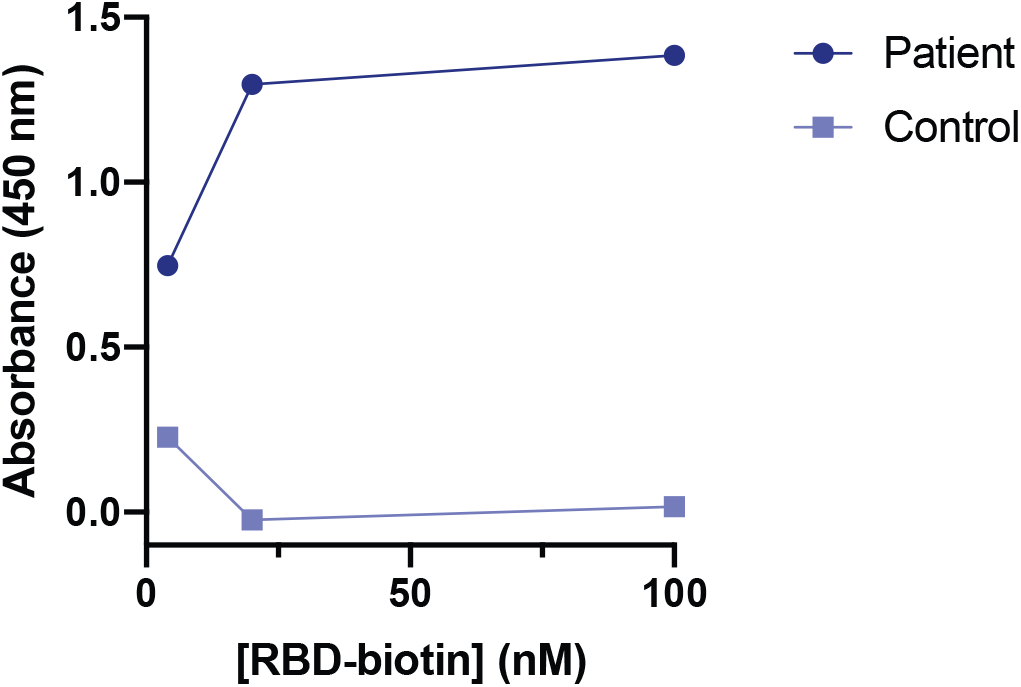
Effect of antigen coating concentration on Protein L-HRP signal. ELISA results from wells coated with varying concentrations of RBD-biotin and incubated with sera from one patient and one healthy control. Sera were diluted 1:50.

**Supplemental Figure 5.**
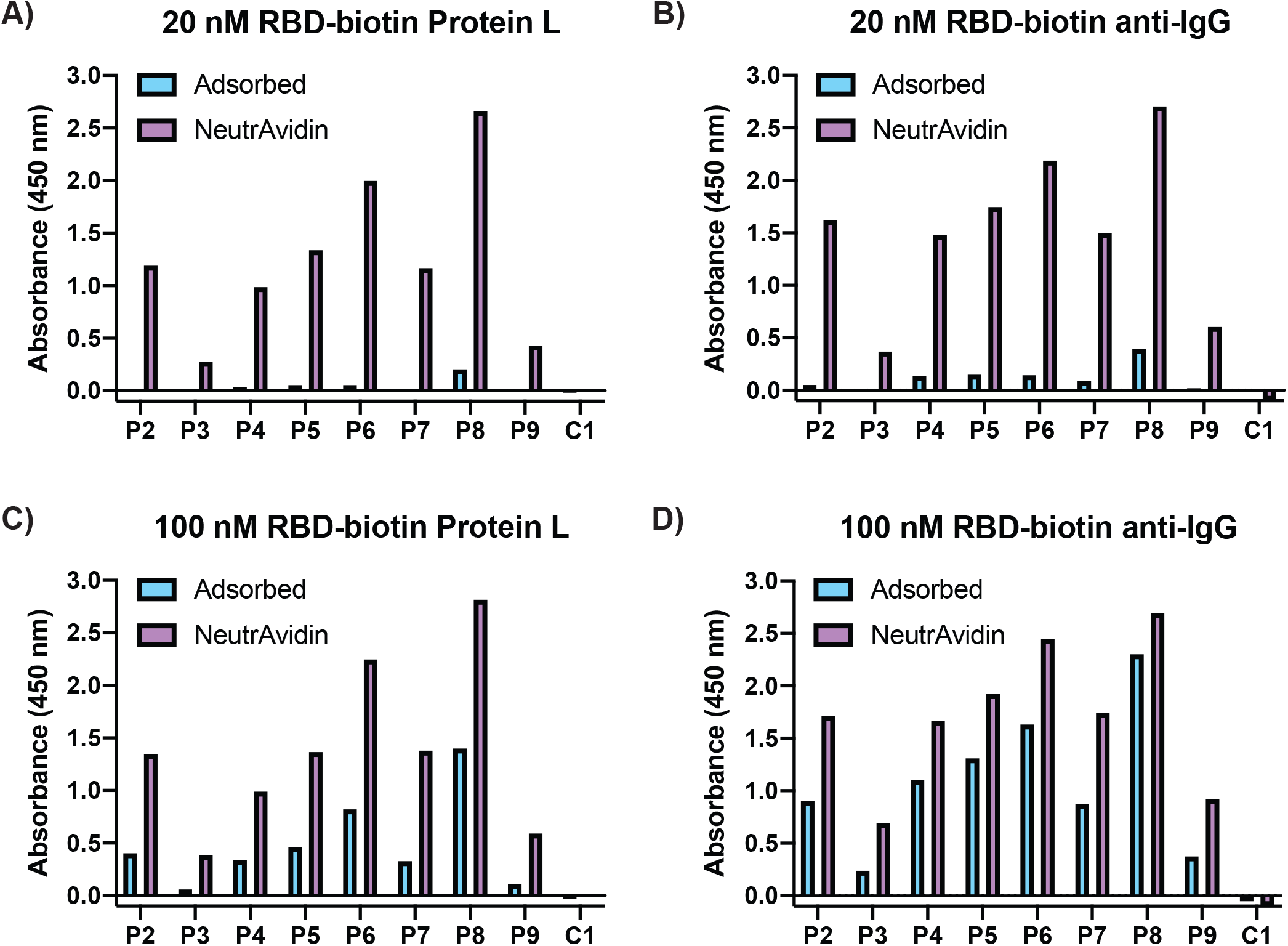
Comparison of passive adsorption and avidin:biotin presentation of RBD-biotin. RBD-biotin was either adsorbed or bound by NeutrAvidin at a concentration of 20 nM (A-B) or 100 nM (C-D) and detected with Protein L-HRP (A, C) or anti-IgG-HRP (B, D). Sera were diluted 1:50.

**Supplemental Figure 6.**
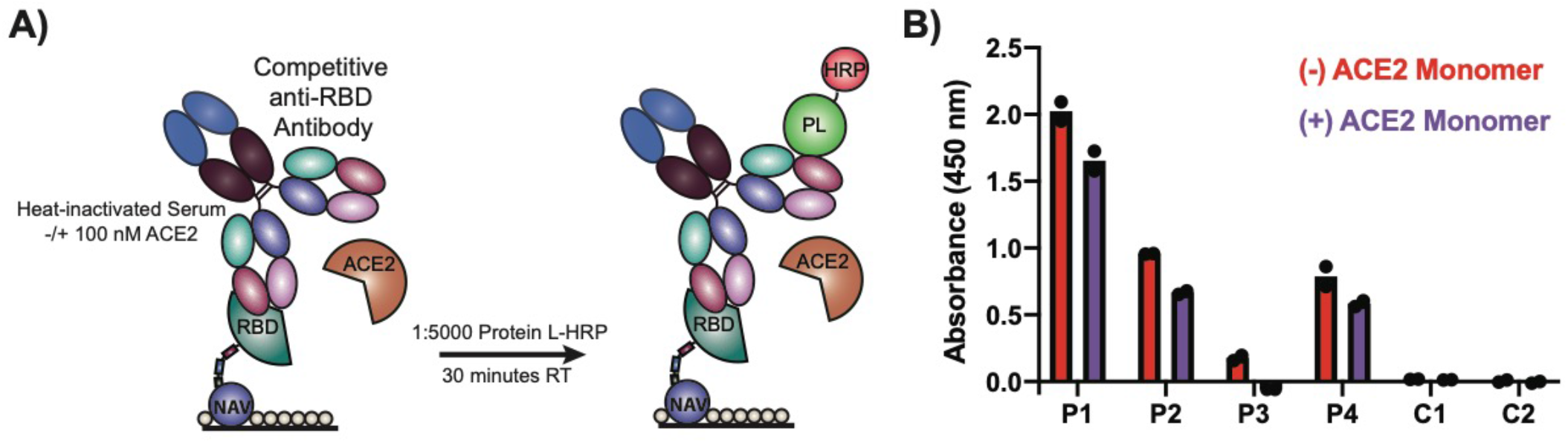
ACE2 monomer only minimally competes with patient anti-RBD antibodies. (A) Schematic of monomeric ACE2 competitive serology ELISA setup. (B) ELISA results from wells coated with 20 nM RBD-biotin and incubated with sera −/+ 100 nM monomeric ACE2 from four patients (P1-P4) and two healthy controls (C1, C2). Sera were diluted 1:50. Dots represent values of technical replicates, bars represent the mean of these replicates.

**Supplemental Figure 7.**
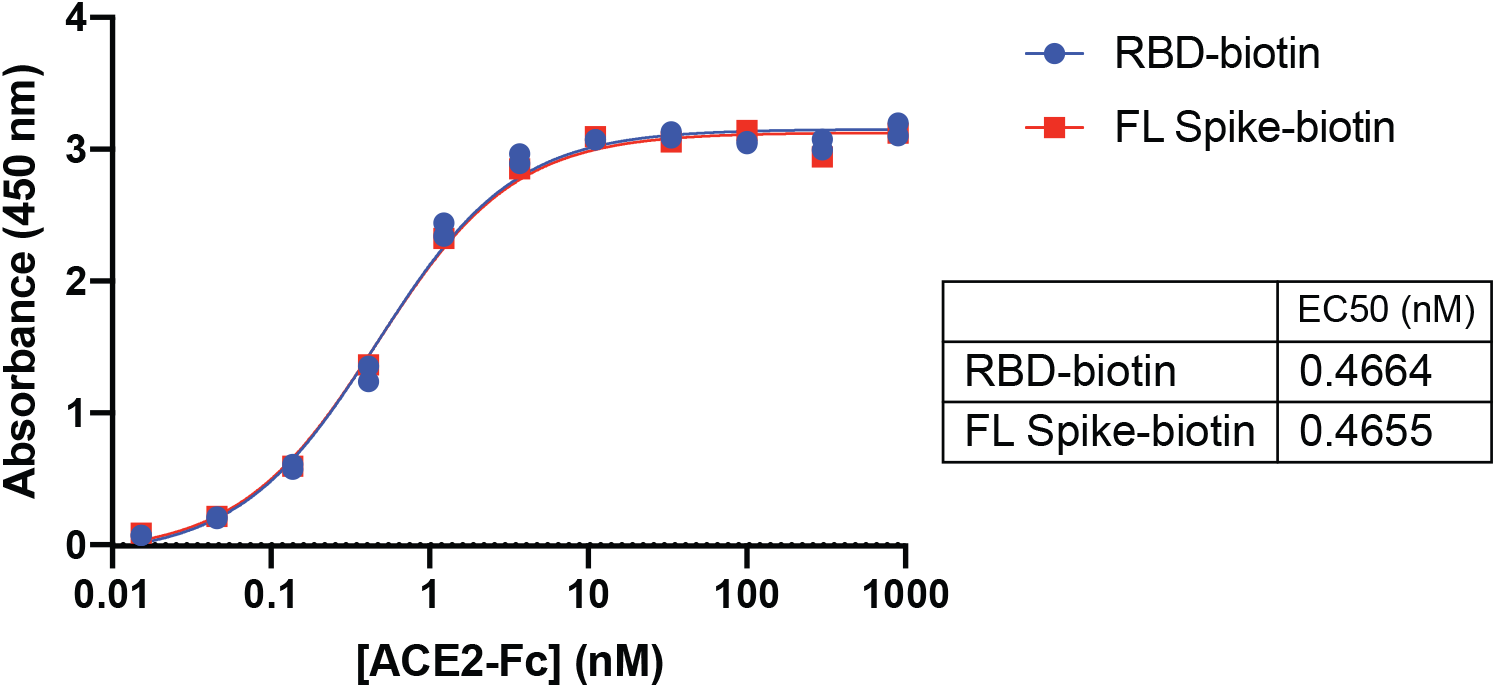
ACE2-Fc saturates plate-bound RBD-biotin and FL Spike-biotin at 100 nM. ACE2-Fc binding was determined using a protocol analogous to the serology ELISA protocol. Instead of serum, three-fold serial dilutions of ACE2-Fc in 1% nonfat milk were incubated in wells coated with NeutrAvidinpresented RBD-biotin or FL Spike-biotin (20 nM coating concentration). ACE2-Fc binding was detected using anti-IgG-HRP. Data are from one experiment.

**Supplemental Figure 8.**
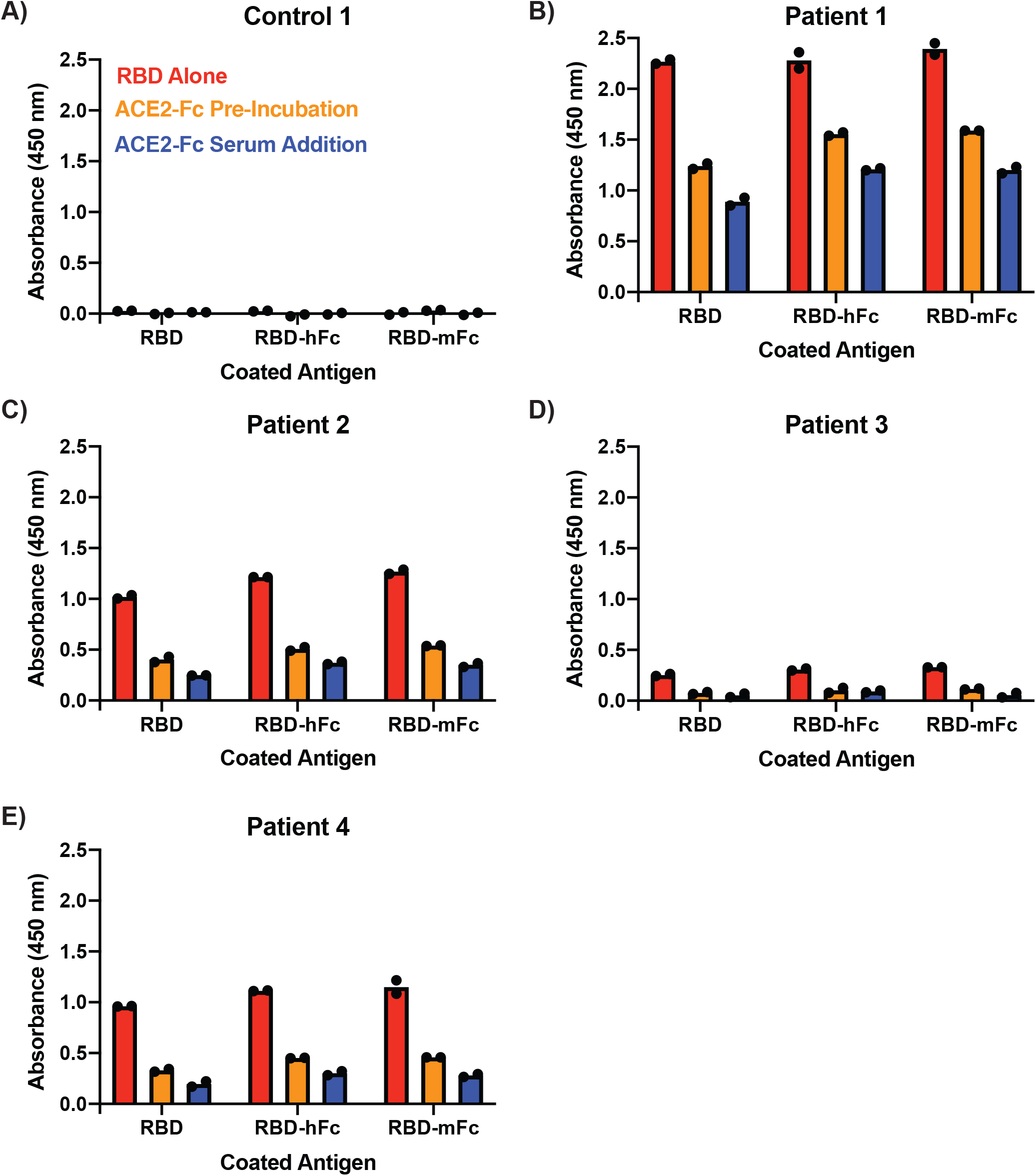
Pretreatment with ACE2-Fc and adding ACE2-Fc to serum yield similar competition profiles. ELISA results from wells coated with 20 nM solutions of the indicated antigens and incubated with sera from four patients and one healthy control. Sera were either added with no ACE2-Fc to untreated wells (red), added to wells pre-treated with 100 nM ACE2-Fc (orange), or supplemented with 100 nM ACE2-Fc (blue) before addition to wells. Sera were diluted 1:50. Dots represent values of technical replicates, bars represent the mean of these replicates.

## REFERENCES

1. Khan, S. et al. Emergence of a Novel Coronavirus, Severe Acute Respiratory Syndrome Coronavirus 2: Biology and Therapeutic Options. Journal of Clinical Microbiology 58, (2020).

2. Wu, F. et al. A new coronavirus associated with human respiratory disease in China. Nature 579, 265–269 (2020).

3. Gralinski, L. E. & Menachery, V. D. Return of the Coronavirus: 2019-nCoV. Viruses 12, 135 (2020).

4. Tai, W. et al. Characterization of the receptor-binding domain (RBD) of 2019 novel coronavirus: implication for development of RBD protein as a viral attachment inhibitor and vaccine. Cellular & Molecular Immunology 1–8 (2020) doi:10.1038/s41423-020-0400-4.

5. Chen, Y., Liu, Q. & Guo, D. Emerging coronaviruses: Genome structure, replication, and pathogenesis. Journal of Medical Virology 92, 418–423 (2020).

6. Wang, C. et al. A human monoclonal antibody blocking SARS-CoV-2 infection. Nature Communications 11, 1–6 (2020).

7. Monteil, V. et al. Inhibition of SARS-CoV-2 Infections in Engineered Human Tissues Using Clinical-Grade Soluble Human ACE2. Cell (2020) doi:10.1016/j.cell.2020.04.004.

8. Lei, C. et al. Potent neutralization of 2019 novel coronavirus by recombinant ACE2-Ig. *bioRxiv* 2020.02.01.929976 (2020) doi:10.1101/2020.02.01.929976.

9. Li, Y. et al. Potential host range of multiple SARS-like coronaviruses and an improved ACE2-Fc variant that is potent against both SARS-CoV-2 and SARS-CoV-1. *bioRxiv* 2020.04.10.032342 (2020) doi:10.1101/2020.04.10.032342.

10. Carter, L. J. et al. Assay Techniques and Test Development for COVID-19 Diagnosis. ACS Cent. Sci. 591–605 (2020) doi:10.1021/acscentsci.0c00501.

11. Stadlbauer, D. et al. SARS-CoV-2 Seroconversion in Humans: A Detailed Protocol for a Serological Assay, Antigen Production, and Test Setup. Current Protocols in Microbiology 57, e100 (2020).

12. Whitman, J. D. et al. Test performance evaluation of SARS-CoV-2 serological assays. *medRxiv* 2020.04.25.20074856 (2020) doi:10.1101/2020.04.25.20074856.

13. Joyner, M. et al. Early Safety Indicators of COVID-19 Convalescent Plasma in 5,000 Patients. *medRxiv* 2020.05.12.20099879 (2020) doi:10.1101/2020.05.12.20099879.

14. Bloch, E. M. et al. Deployment of convalescent plasma for the prevention and treatment of COVID-19. J Clin Invest (2020) doi:10.1172/JCI138745.

15. Amanat, F. et al. A serological assay to detect SARS-CoV-2 seroconversion in humans. Nature Medicine 1–4 (2020) doi:10.1038/s41591-020-0913-5.

16. Lui, I. et al. Trimeric SARS-CoV-2 Spike interacts with dimeric ACE2 with limited intra-Spike avidity. *bioRxiv* 2020.05.21.109157 (2020) doi:10.1101/2020.05.21.109157.

17. Hornsby, M. et al. A High Through-put Platform for Recombinant Antibodies to Folded Proteins. Mol Cell Proteomics 14, 2833–2847 (2015).

18. Chen, X. et al. Human monoclonal antibodies block the binding of SARS-CoV-2 spike protein to angiotensin converting enzyme 2 receptor. Cellular & Molecular Immunology 1–3 (2020) doi:10.1038/s41423-020-0426-7.

19. Chi, X. et al. A potent neutralizing human antibody reveals the N-terminal domain of the Spike protein of SARS-CoV-2 as a site of vulnerability. *bioRxiv* 2020.05.08.083964 (2020) doi:10.1101/2020.05.08.083964.

20. To, K. K.-W. et al. Temporal profiles of viral load in posterior oropharyngeal saliva samples and serum antibody responses during infection by SARS-CoV-2: an observational cohort study. The Lancet Infectious Diseases 20, 565–574 (2020).

21. Wu, F. et al. Neutralizing antibody responses to SARS-CoV-2 in a COVID-19 recovered patient cohort and their implications. *medRxiv* 2020.03.30.20047365 (2020) doi:10.1101/2020.03.30.20047365.

22. He, Y., Zhou, Y., Siddiqui, P. & Jiang, S. Inactivated SARS-CoV vaccine elicits high titers of spike protein-specific antibodies that block receptor binding and virus entry. Biochemical and Biophysical Research Communications 325, 445–452 (2004).

23. Berry, J. D. et al. Neutralizing epitopes of the SARS-CoV S-protein cluster independent of repertoire, antigen structure or mAb technology. mAbs 2, 53–66 (2010).

